# Phenotypic convergence: a novel phenomenon in the diagnostic process of Mendelian genetic disorders

**DOI:** 10.1101/2023.01.17.23284691

**Authors:** Rory J Tinker, Josh Peterson, Lisa Bastarache

## Abstract

**Introduction:** The study of Mendelian disease has yielded a large body of knowledge about the phenotypic presentation of disease. Less is known about the way the diseases are reflected in the electronic health record (EHR).

**Aim:** To develop an EHR-based model of the diagnostic trajectory and investigate data availability and the longitudinal distribution of signs and symptoms of a Mendelian disorder within EHRs.

**Methods:** We created a conceptual model to specify key time points of the diagnostic trajectory and applied it to individuals with genetically confirmed hereditary connective tissue diseases (HCTD). Using the model, we assessed EHR data availability within each time interval. We tested the performance of phenotype risk scores (PheRS), an algorithm that detects Mendelian disease patterns and assessed the phenotypic expression of HCTD over the diagnostic trajectory.

**Results:** We identified 251 individuals with HCTD; 79 (35%) of these patients had a fully ascertained diagnostic trajectory. There were few documented signs and symptoms prior to clinical suspicion that evoked an HCTD disorder (median PheRS 0.14); once suspicion was documented, median PheRS increased to 1.87 (SD). The majority (72%) of phenotypic features were identified post clinical suspicion.

**Discussion:** Using a novel conceptual model for the diagnostic trajectory of Mendelian disease, we demonstrated that phenotype ascertainment is, in part, driven by the diagnostic process and that many findings are only documented following clinical suspicion and diagnosis, a process we term phenotypic convergence. Therefore, algorithms that aim to detect undiagnosed Mendelian disease should censor EHR data to avoid data leakage.

## Introduction

The study of Mendelian disease has yielded a great body of knowledge about the phenotypic manifestations of Mendelian disease. ^1–4^ This knowledge has been generated through the careful study of affected individuals which have been summarized in resources like The Online Mendelian Inheritance in Man (OMIM). OMIM currently has detailed clinical descriptions for thousands of genetic diseases and is an essential resource for recognizing and diagnosing genetic disease.

Despite the availability of sophisticated knowledgebases, recognizing genetic disease can still be a great challenge for clinicians, leading to prolonged diagnostic delay (DD) ^5–7^. DD continues to be a problem for a variety of genetic diseases that has not consistently improved with time ^8–12^. Current evidence suggests that neither increased availability of genetic testing nor targeted awareness campaigns aimed at increasing awareness of rare diseases are sufficient to fully addressing DD ^10–12^. The persistent problem has led to an interest in diagnosis support systems to identify undiagnosed patients using data from the electronic health record (EHR). ^13–17^.

To effectively identify undiagnosed patients, EHR-based algorithms must “recognize” the clinical patterns of the genetic disease, just as clinicians do. But unlike clinicians, these algorithms cannot examine patients. Rather, they must make use of phenotypic clues that are available in real world clinical data. As such, they must address two key challenges of EHR data ^18,19^. First, EHRs are often missing data. A leading cause of incompleteness is due to information fragmentation, which occurs when patients receive care at multiple care sites.^20^ EHR chart fragmentation may be particularly problematic for patients with rare genetic disease, who are more likely to seek care at disparate referral centers. For algorithms that seek to identify undiagnosed disease, fragmented EHRs may be inappropriate for training and testing. Secondly, EHRs are affected by ascertainment bias. EHRs contain myriad observations, measurements, and diagnoses that simultaneously reflect the physiology of a patient as well as the process of healthcare itself. True facts about a patient are often missing from a record until they become clinically relevant.

In this paper, we study the phenotypic manifestations of genetic disease from the perspective of the EHR. A recent scoping review noted that there is no systematic framework to test, train, and evaluate EHR-based diagnosis support systems in rare disease, hindering the ability to realistically assess and compare different algorithms whilst increasing the risk of data leakage ^22^. Our conceptual model seeks to describe the diagnostic trajectory as reflected in the EHR. The notion of a diagnostic trajectory is analogous to a disease trajectory, in which the progression of a disease is tracked over time. ^23^ The model defines key timepoints in the diagnostic process and enables the assessment of data availability and at different phases of the diagnostic trajectory. Applying this framework to hereditary connective tissue diseases (HCTDs), we (1) examine the effect of EHR fragmentation on the feasibility of a diagnosis support system at an academic medical center (2) measure EHR data availability within each diagnostic time interval defined by clinical suspicion or diagnosis, and (3) characterize the way ascertainment bias influences how diseases are documented in the EHR data at different points. We find that EHR fragmentation is a major barrier to ascertaining undiagnosed patients prior to clinical suspicion for disease. And we note that the ascertainment of key phenotypes often occurs only after clinical suspicion and/or diagnosis of a disease. We term this concept phenotypic convergence whereby the phenotypic EHR arises after clinical suspicion of the diagnosis.

## Material and methods

### Defining the conceptual model and keys terms

We defined a conceptual model (Figure 1) that describes key events of the diagnostic process as reflected in EHR data, including: first encounter within the health system, first clinical suspicion (the date when a disease is first mentioned in the record), diagnosis (the date when a clinical diagnosis is established), and final encounter. These events delineate the following time intervals: Pre-ascertainment, Pre-suspicion, Pre-diagnosis, Suspicion to diagnosis and post-diagnosis. Within these intervals, we characterize data availability by both quantity (number of encounters) and longitudinally (length of time in days). A patient has a fully ascertained diagnostic trajectory if they have encounters in the pre-suspicion, suspicion to diagnosis, and post-diagnosis intervals. A full glossary of the definition of terms can be found in Table 1.

**Table 1:** A glossary of key terms used in the current study.

| Term | Definition | Note |
| --- | --- | --- |
| Date of birth | Date of patient’s birth as listed in the demographics table | - |
| Date at first encounter | The first encounter date recorded in an EHR | Encounter dates can be defined in several ways (e.g. billing codes, clinic notes). For a predictive algorithm, encounter should be defined as dates where new phenotypic information is ascertained (i.e. an ascertainment event). For algorithms that use claims data to define features, the ascertainment event may be defined as billing days. For those that use clinical notes, a clinical encounter may be used. |
| Date of first clinical suspicion | The first date a clinician documents a suspicion of a disease in clinical notes, whether specific (e.g. “Marfan syndrome”) or more general (e.g. “connective tissue disease”) | - |
| Date of diagnosis | The first date when a treating physician states that the patient is clinically diagnosed with the condition. This declaration is often preceded by statements that the diagnosis is “probable” or “likely.” Such conditional statements do not qualify as a diagnosis | - |
| Date at last encounter | Last encounter date in the EHR | - |
| Pre-ascertainment period | Birth to first encounter in calendar days | - |
| EHR capture period | Date of clinical date of first encounter to date of last encounter |  |
| Pre-suspicion interval | First encounter to one week prior to clinical suspicion in calendar days | - |
| Suspicion to diagnosis interval | Date of suspicion to one week prior to diagnosis |  |
| Pre-diagnosis interval | First encounter to one week prior to diagnosis in calendar days | - |
| Post-diagnosis interval | Date of diagnosis to final encounter in calendar days | - |
| Data Quantity | The number of unique encounter dates | The pre-suspicion data quantity indicates the amount of information available on which to make a prediction |
| Data longitudinally | Length of time interval in days | The longitudinally of the pre-suspicion interval defines a theoretical maximum of how much earlier a diagnosis could have been made. |

**Figure 1:**
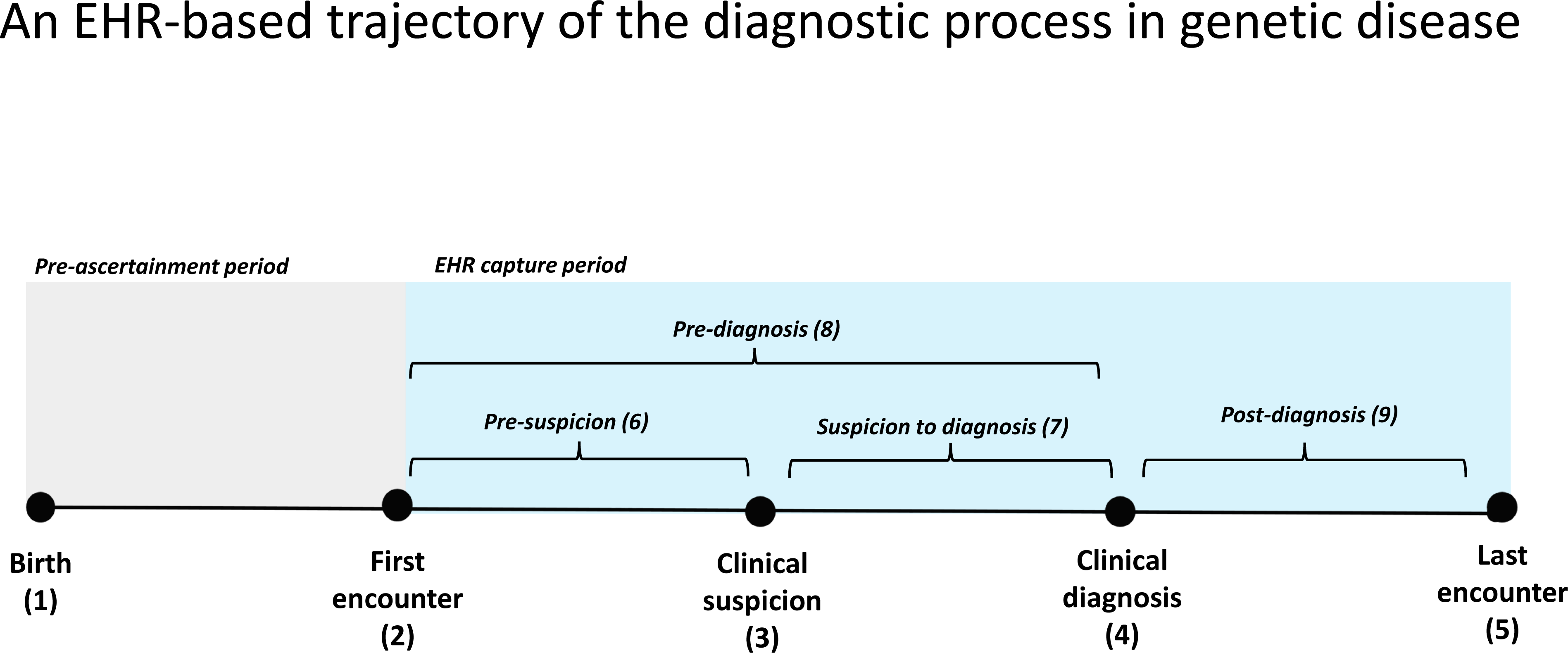
A graphical representation of the diagnostic trajectory as represented in EHR data. This model is intended to help assemble and code EHR data to both study the diagnostic process and test/train models to identify undiagnosed patients. It includes the following temporal events: (1) Date of birth, (2) Date at first encounter, (3) Date of first clinical suspicion, (4) Date of clinical diagnosis and (5) Date at last encounter. These timepoints define the following longitudinal intervals: (6) Pre-suspicion interval (first encounter to one week prior to clinical suspicion), (7) Suspicion to diagnosis interval (date of suspicion to one week prior to diagnosis), (8) Pre-diagnosis interval (first encounter to one week prior to diagnosis) and (9) post-diagnosis interval (date of diagnosis to final encounter). Two periods were also defined (grey) Pre-ascertainment period (birth to first encounter) and (blue) EHR capture period (date of clinical date of first encounter to date of last encounter). Key timepoints are indicated by black circles, key time intervals are denoted by brackets and key periods are indicated in color.

### Data source

Our cohort comprised all individuals with at least three encounters within Vanderbilt University Medical Center (VUMC) between 2002-01-01 and 2022-01-01. Data was drawn from the research derivative (RD), a copy of VUMC’s EHR stored in the Observational Medical Outcomes Partnership (OMOP) common data model that includes demographics, notes, International Classification of Disease (ICD) and Current Procedural Terminology (CPT) codes used in this project ^24^.

### Applying the conceptual model to HCTDs

We elected to study HCTD, a class of diseases with prominent multi-system characterized by phenotypes related to the musculoskeletal system, vision, and (with the exception of STL) cardiovascular system.^25–27^ We included the most prevalent HCTDs: Marfan syndrome (MFS), Loeys-Dietz syndrome (LDS), Sticker syndrome (STL), and Ehlers Danlos syndrome (EDS). We used OMIM to generate a list of causal genes in the phenotypic series for LDS (PS609192), STL (PS108300), EDS (PS130000), and MFS (OMIM# 154700). We indexed all clinical notes for HCTD gene names. *DSE* (Dermatan Sulfate Epimerase) was excluded due to excessive false positive mentions. The chart of every individual with a mention of a gene name in their record was manually reviewed for confirmation, and individuals with a pathogenic, diagnostic variant were flagged for further review.

To define the diagnostic trajectory of each HCTD patient, we extracted key timepoints from the EHR. (Figure 1). Date of birth was derived from the demographics table, and the first and last encounters were defined using the first and last ICD billing code dates. Initial suspicion was defined as the first mention of an HCTD in clinical notes, either by a specific disease name (e.g. Marfan syndrome) or disease class (e.g. connective tissue disease), and the date of diagnosis was defined as the date that the diagnosis was established; both were defined via manual chart review. We calculated the age of the HCTD patients at each of the key timepoints and computed longitudinally (length of time in days) and quantity within each interval. Encounters were defined using ICD billing dates.

### Phenotype risk score analysis

Phenotype risk scores (PheRS) is a measurement of the similarity of a patient’s EHR data (specifically ICD codes) and the clinical presentation of a disease as described in OMIM. PheRS represents characteristic phenotypes of genetic disease with phecodes: ICD based high-throughput phenotype. PheRS has been used to assess the pathogenicity of genetic variants as well as to detect undiagnosed individuals using EHR data.^28,29^ Here we tested the ability of PheRS to distinguish between HCTD cases and controls at multiple time points along the diagnostic trajectory.

As OMIM has multiple clinical synopses for LDS, STL, and EDS (one for each causal gene), we merged the synopses into a single feature set for each disease. We applied PheRS for each of the four HCTDs to our entire cohort of 1.8 million individuals. Regression was used to normalize the PheRS by gender, age, and record length, producing a “residualized PheRS” (rPheRS). We compared rPheRS in cases versus controls using the Wilcox rank sum test. We also counted the number of cases with “highly elevated” scores (rPheRS > 4 standard deviations above the mean).

We repeated the above analysis after censoring HCTD data at different timepoints of the diagnostic trajectory, using ICDs from the pre-suspicion interval, the pre-diagnosis interval, and the post-diagnosis interval. For each analysis, a new cohort was generated in which the data from the HCTD patients was censored to match the target interval and with covariates that reflected the censored data.

### Individual phenotype analysis

We explored the phenotypes driving the changes in rPheRS across the diagnostic trajectory by analyzing when individual HCTD phenotypes appear in the EHR. For each HCTD feature, we counted the number of individuals who had a phecode dated prior to suspicion, between suspicion and diagnosis, following diagnosis, and ever. We repeated this analysis after grouping phenotypes into phecode categories.

### Ascertainment analysis

We extracted CPT codes and dates for the following procedures: Transthoracic echocardiography (93303, 93304, 93306, 93307, 93308); ophthalmological examination and evaluation (92002, 92004, 92012, 92014, 92015, 92018, 92019); and complete blood count (85027, 85025). We counted the number of individuals within each code group as well as the total number of procedure codes prior to suspicion. We reviewed the charts of individuals with aortic aneurysms to determine the first date this phenotype was mentioned in the chart and compared that date to the first billing code date.

### Statistical tools

All statistical analyses were conducted in R. We used the phers R package to generate scores^30^. For the residual scores (rPheRS), we included covariates for gender, age at first visit (days), age at last visit (days), and total number of ascertainment events (unique dates with ICD codes). PhecodeX was used to translate ICDs into phecodes ^31^. Our list of HCTDs

## Results

We indexed the records of 1.8 million patients for 32 HCTD genes. After excluding *DSE*, a total of 2127 charts mentioned one or more of these genes. Through manual review, we identified 253 individuals with a genetically confirmed diagnosis of HCTD for 13 distinct OMIM IDs. (Table S1).

### The impact of data fragmentation on the ascertainment of a complete diagnostic trajectory

Among the HCTD patients, 77 (30%) were diagnosed prior to their first visit to VUMC. For another 89 individuals (35%), the date of clinical suspicion occurred within a week of their first visit. In total, 88 individuals (34%) were found to have fully ascertained diagnostic trajectories, including 33 MFS, 19 EDS, 17 LDS and 18 STL cases. (Table 2).

**Table 2:** EHR data availability in HCTD cohort. Abbreviations MFS - Marfan syndrome; EDS - Ehlers-Danlos syndromes; LDS - Loeys-Dietz syndrome; STL - Stickler syndrome; ALL – All Hereditary connective disease

|  | MFS | EDS | LDS | STL | ALL |
| --- | --- | --- | --- | --- | --- |
| Diagnosed before first visit |  |  |  |  |  |
| N (%) | 55 (37.9) | 7 (19.4) | 7 (21.9) | 8 (20) | 77 (30.4) |
| Suspicion on first encounter | 57 (39.3) | 10 (27.7) | 8 (25.0) | 14 (35) | 89 (35.2) |
| Fully ascertained trajectory | 33 (22.8) | 19 (52.8) | 17 (53.1) | 18 (45) | 87 (34.4) |
| Total | 145 | 36 | 32 | 40 | 253 |

Among individuals with fully ascertained trajectories, the age of diagnosis varied widely, with a median of 13 (Q1 – Q3: 4.6-28.2) Figure 2A). 30 patients (34%) were diagnosed as adults, which is suggestive of a long diagnostic delay. (Table 3)

**Table 3:**
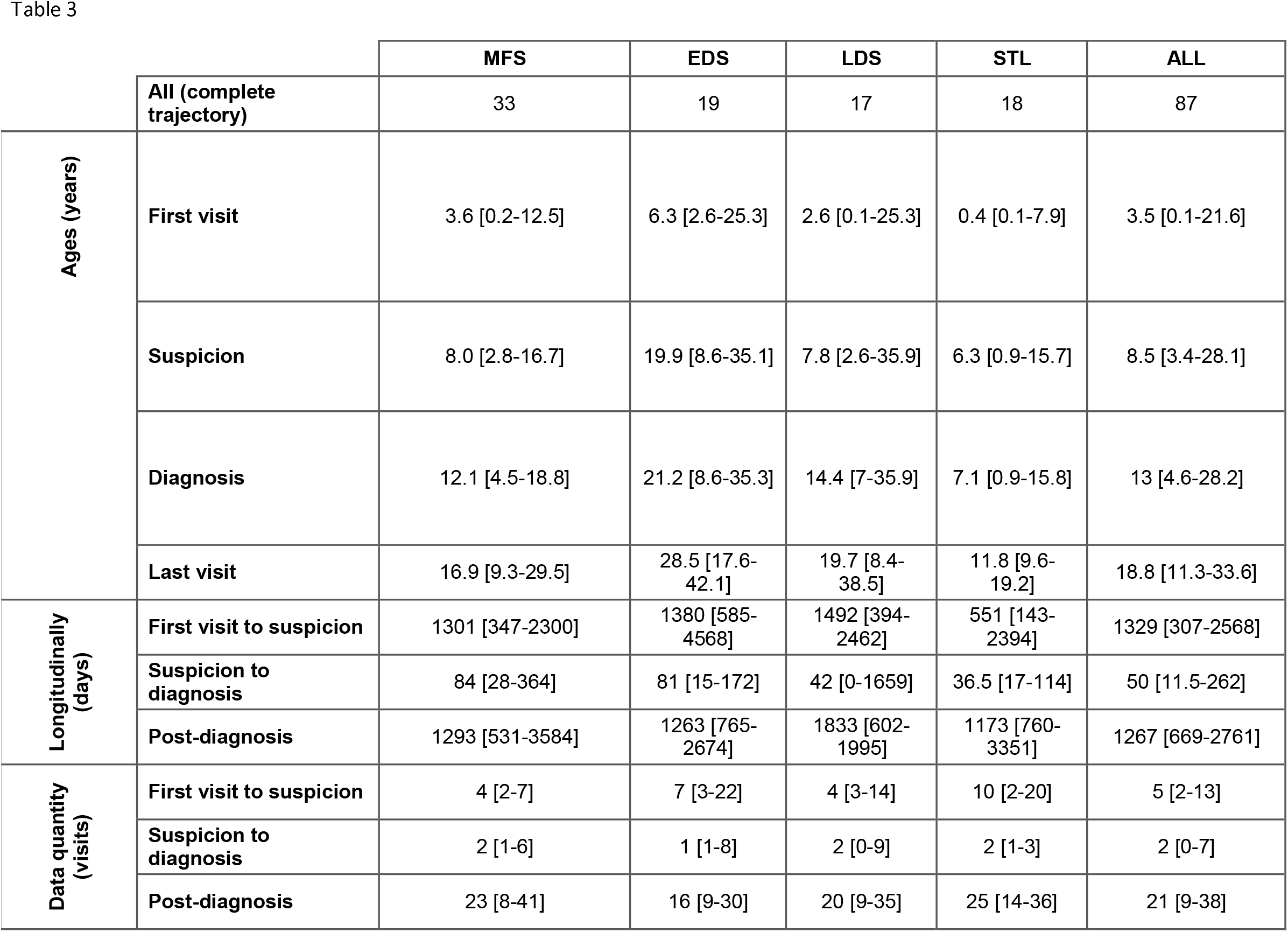
The key temporal points and data availability of HCTD individuals with fully ascertained trajectories. **Abbreviations** MFS - Marfan syndrome; EDS - Ehlers-Danlos syndromes; LDS - Loeys-Dietz syndrome; STL - Stickler syndrome; ALL – All Hereditary connective disease

**Figure 2:**
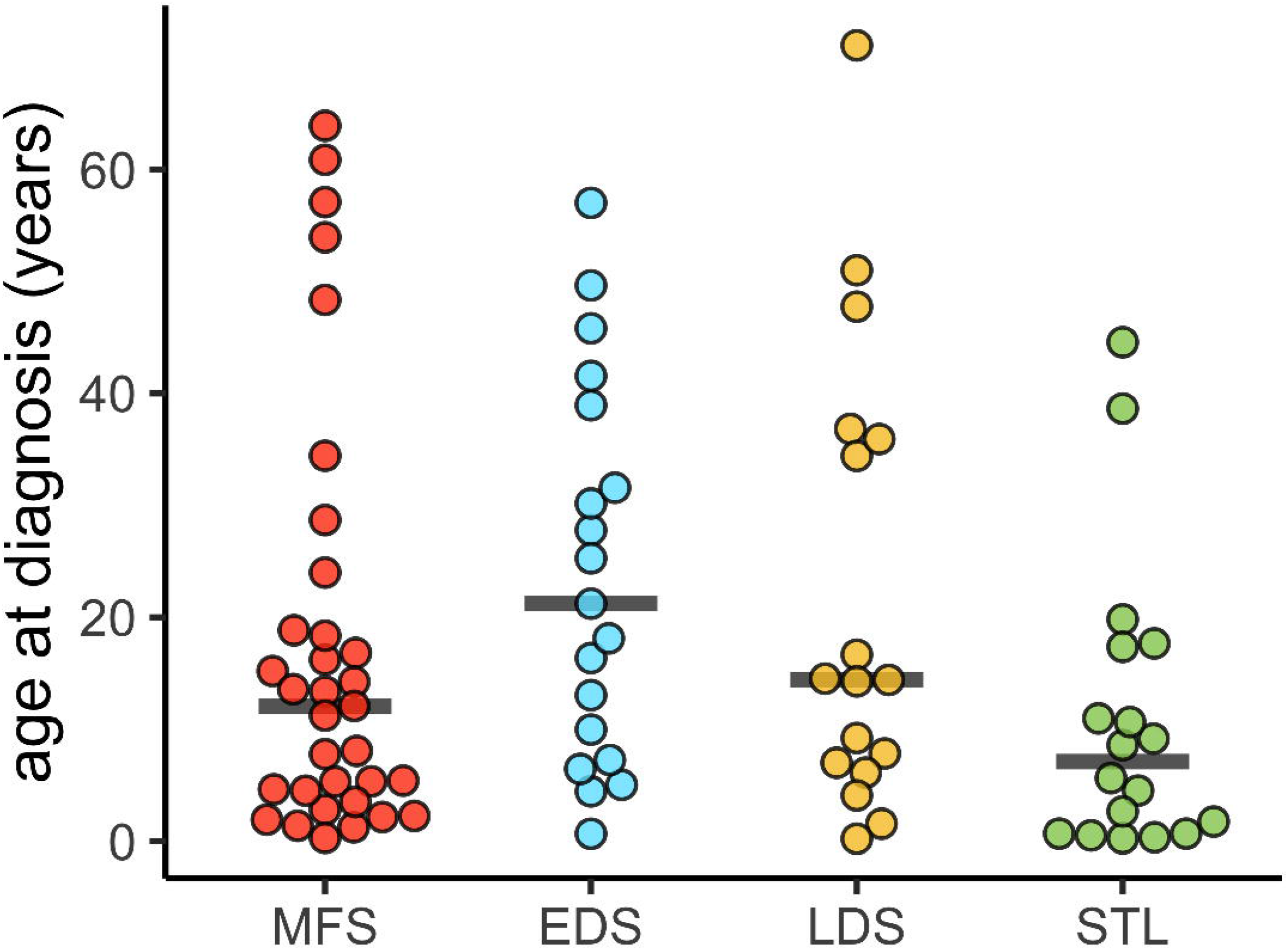
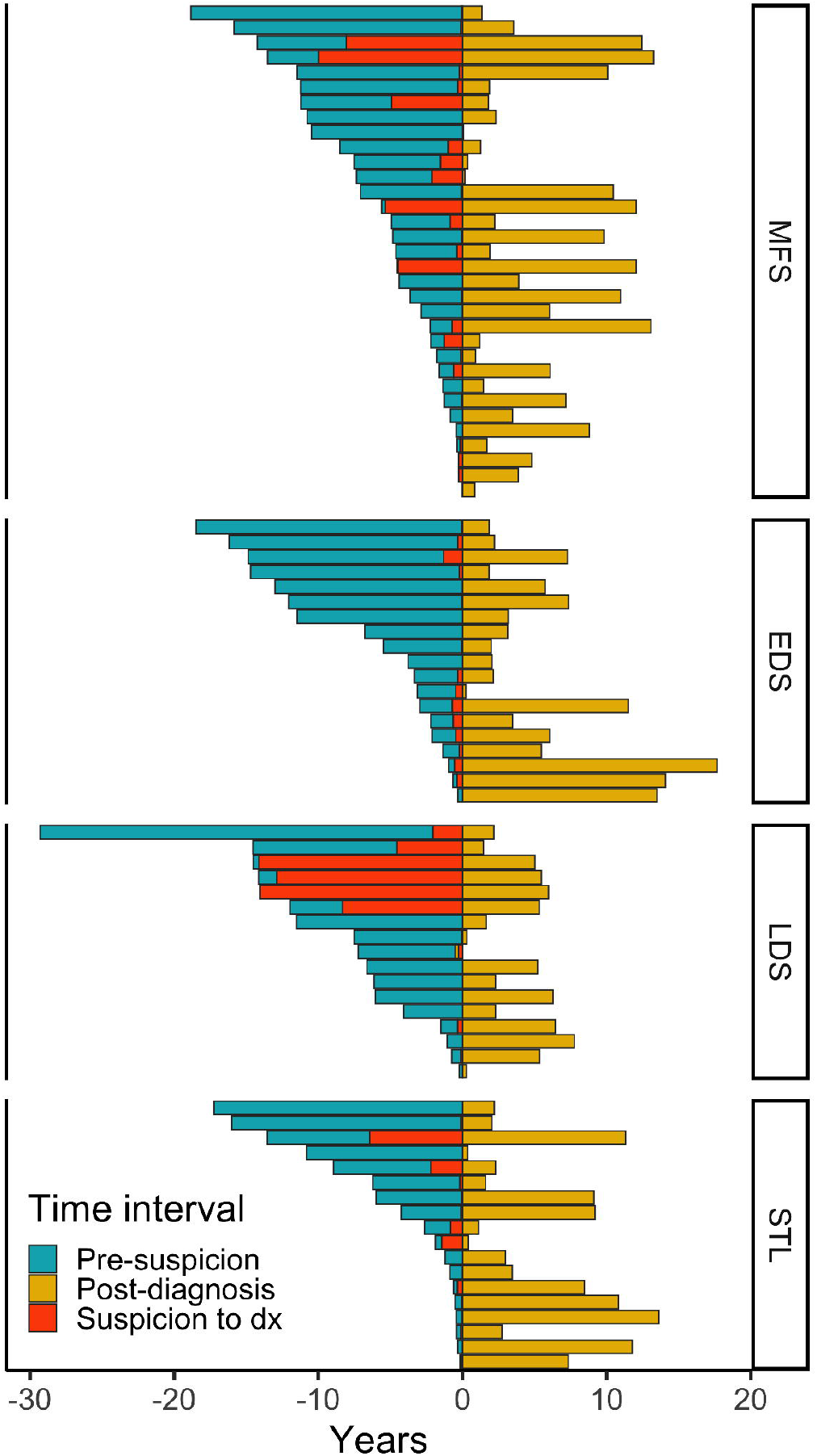
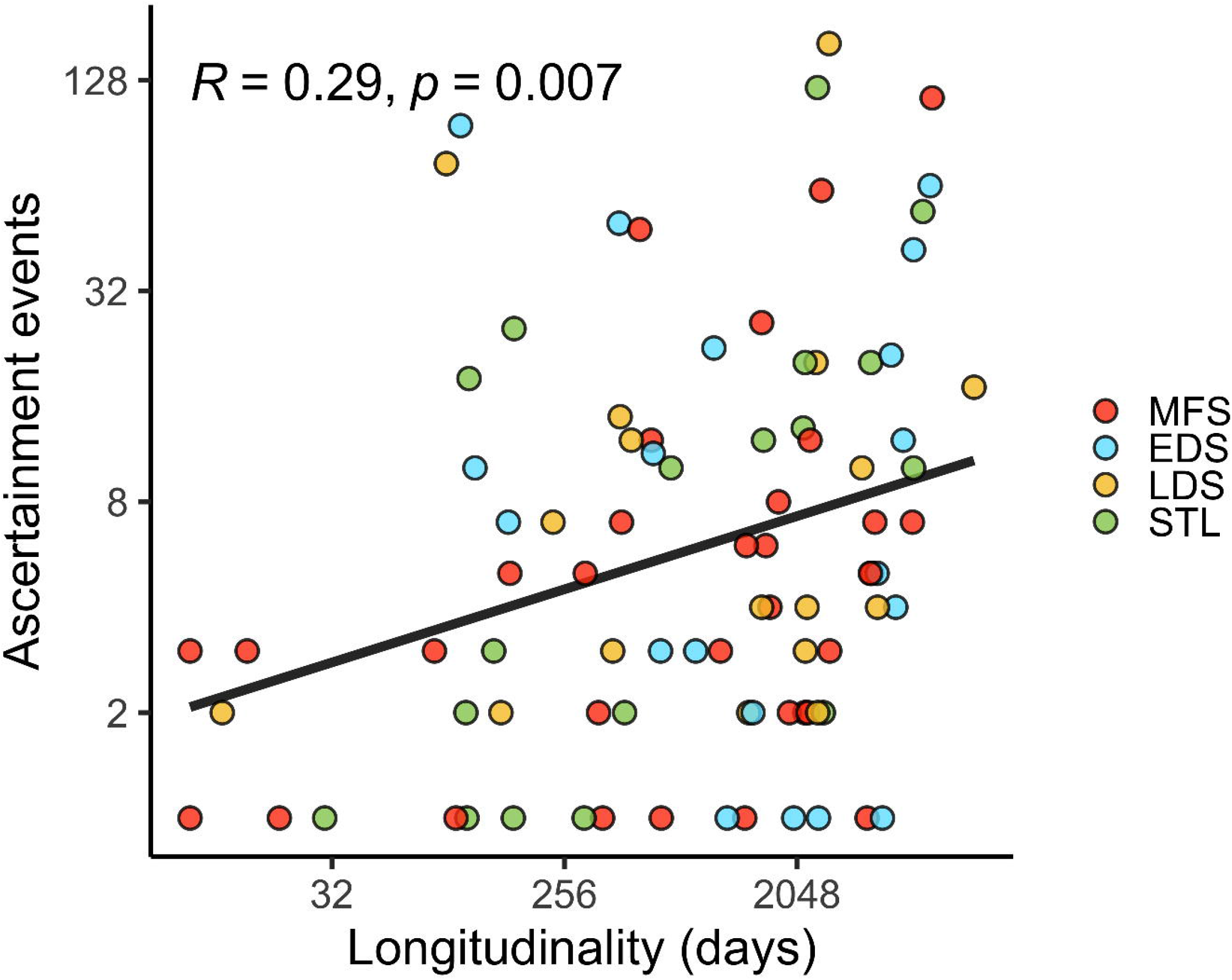
**2A**. Age of diagnosis for all individuals in the HCTD cohort. Each dot represents an individual patient, color coded by HCTD. **2B**. Longitudinally of EHR at different time intervals. Each patient is represented as a bar. Time prior to diagnosis is plotted in the negative direction and post-diagnosis in the positive direction. **2C**. Correlation of longitudinally and data quantity. Each dot represents an individual patient, color coded by HCTD. The plot is scaled by log2 for readability. R is for Pearson correlation

An algorithm designed to shorten diagnostic delay must rely on data from before the date of diagnosis or, better yet, clinical suspicion. Thus, the measures of longitudinally and quantity indicate how much information is available for predictive algorithms. The median time from first visit to clinical suspicion was 3.6 years [307 days – 7.0. years], indicating a theoretical opportunity to shorten the time to diagnosis. Within that interval, a median of 5 visits [2 – 13] occurred. (Figure 2B) Longitudinally and quantity were significantly correlated (R=0.3; p=0.005), though there were many instances with high data quantity and low longitudinally. (Figure 2C) The median time from clinical suspicion to diagnosis was 50 days [11.5-262], indicating that, for most patients, diagnosis occurred relatively quickly once a HCTD was suspected.

### PheRS signal increases over the diagnostic trajectory

The PheRS feature definition included a total of 172 phenotypes (Table S2). When all available EHR data was used, we found that the PheRS of HCTD patients had significantly higher scores than controls for each disease (p <4×10^−8^) PheRS remained significantly elevated in cases versus controls for all time periods tested, but the degree of separation differed. The median rPheRS was 0.14 [-0.03 - 1.48] in the pre-suspicion interval, 1.36 [-0.02 - 3.47] in the pre-diagnosis interval, and (Figure 3; Table S3). These differences were significant in the paired Wilcox rank sum test (p<0.05), including the difference between the pre-suspicion and post-diagnosis intervals (1.87 [0.86 - 3.91]; p=3.1×10^−7^) despite having similar longitudinality in both. (Table S4). Additionally, the number of individuals with highly elevated scores increased across the diagnostic trajectory, from 6 (6.9%) in the pre-suspicion interval, to 16 (18%) in the pre-diagnosis interval, to 21 (24%) in the post-diagnosis interval, and 30 (34%) when all available data was used. (Table S5)

**Figure 3:**
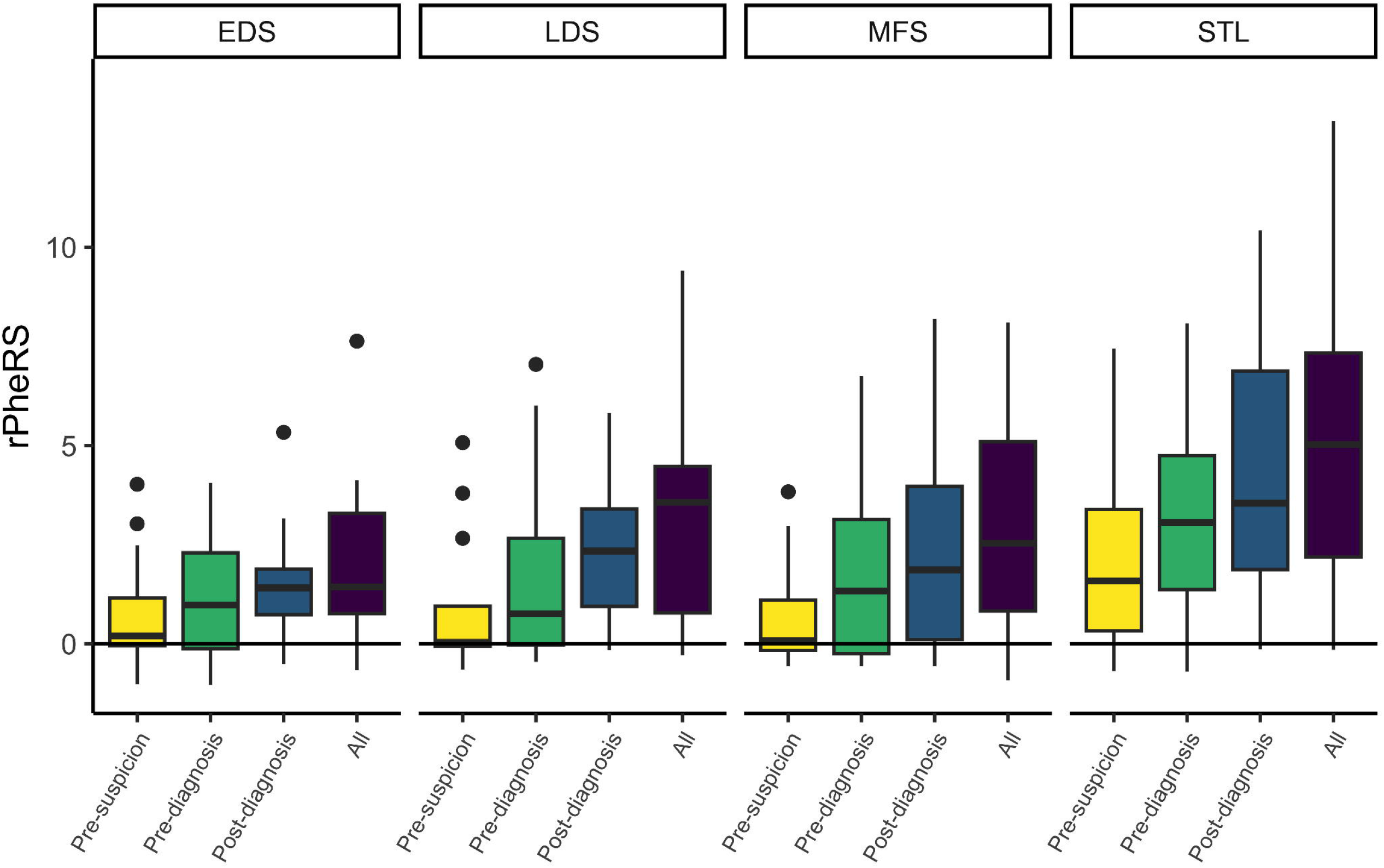
Changes in the PheRS throughout the diagnostic process of HCTD’s (MFS=33; EDS=19; LDS=17; STL=18). There were statistically significant increases in PheRS scores before suspicion to before diagnosis before diagnosis to after diagnosis for all HCTD’s. Abbreviations MFS - Marfan syndrome; EDS - Ehlers-Danlos syndromes; LDS - Loeys-Dietz syndrome; STL - Stickler syndrome; ALL – All Hereditary connective disease

### Phenotypic features of HCTD are more likely to be present in the EHR after clinical suspicion

Only a minority of HCTD-related phenotypes were first ascertained after clinical suspicion (Figure 4a). This pattern was true for most individual phenotypes. For example, for MFS patients only three features were most likely to be ascertained prior to suspicion (Tall stature, Congenital deformities of skull, face, and jaw, and Hammer toe), while 11 were more common during/after diagnosis, including aortic aneurysm and ectasia, mitral valve disorders, pectus excavatum, and lens dislocation. Similar patterns were found for the other three HCTDs (Tables S6-9).

**Figure 4:**
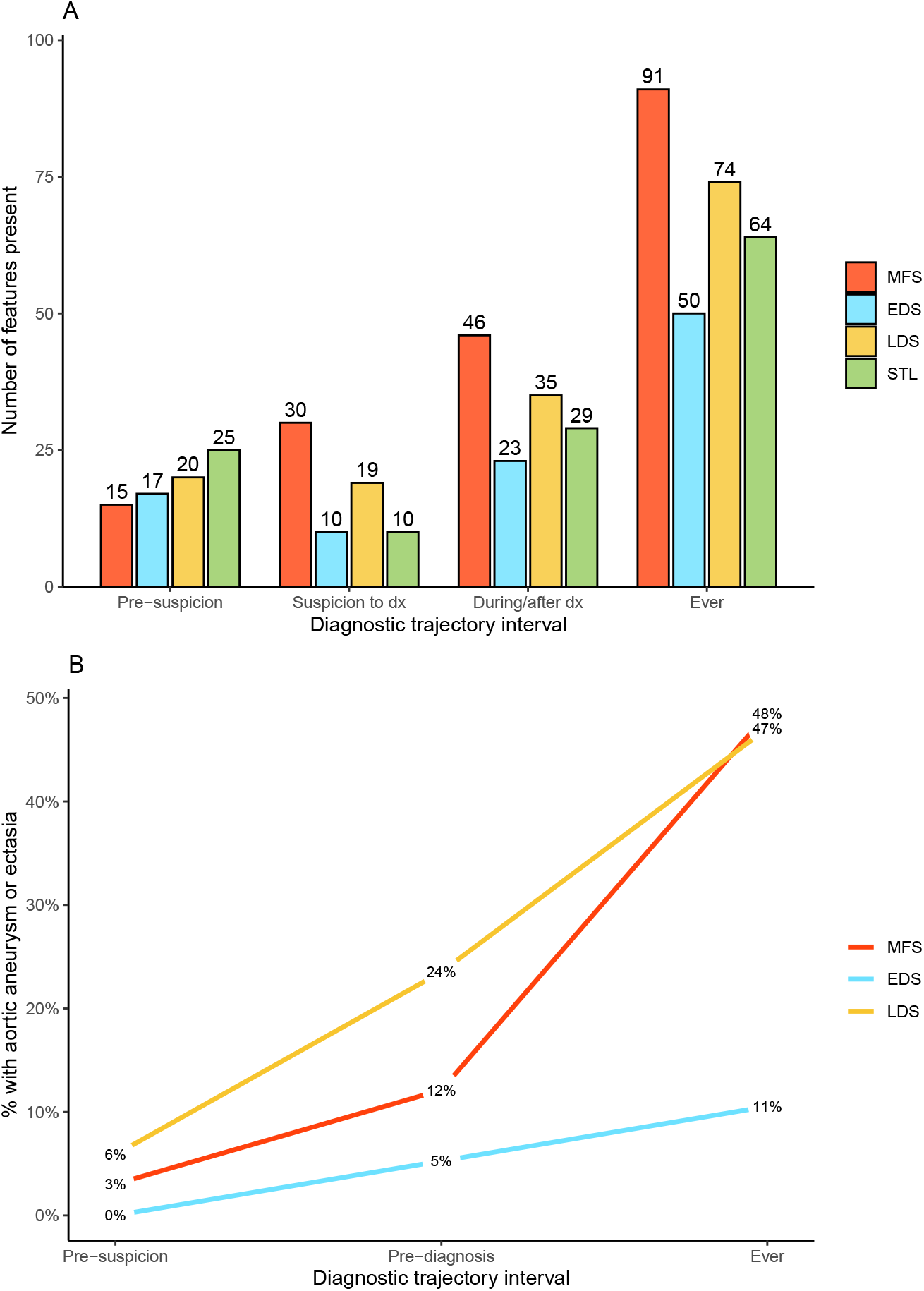
**4a** A bar chart showing the timing of HCTD phenotype ascertainment. The number above each bar represents the percentage of HCTD phenotypes that were first ascertained in that time interval. Among the 91 total MFS phenotypes, 16% were first ascertained before suspicion, 33% between suspicion and diagnosis, and 51% during or after diagnosis. For all diseases, the majority of related phenotypes were first ascertained following clinical suspicion for an HCTD. **4b**. Cumulative prevalence of aortic aneurysm and ectasia among MFS, EDS, and STL patients (LDS is not included because AA is not a characteristic feature for this disease). The percentage of patients with a phecodes for AA increases across the diagnostic trajectory. Abbreviations MFS - Marfan syndrome; EDS - Ehlers-Danlos syndromes; LDS - Loeys-Dietz syndrome; STL - Stickler syndrome; ALL – All Hereditary connective disease

Aortic aneurysm (AA) is highly specific feature of most HCTDs which on its own should raise clinical suspicion and was also the most common overall phenotypic feature present in our cohort, seen in 30% of individuals ^32^. The cumulative prevalence of AA increased substantially after suspicion and diagnosis (Figure 4B).

### Investigating ascertainment procedures, age related expressivity, and phenotyping error in EHR phenotypes

We hypothesized that phenotypic convergence occurs because suspicion and diagnosis prompts the clinician to look for and ascertain signs and symptoms of a genetic disease. Thus, phenotypes are newly ascertained only after a disease is suspected, even if they were present in the patient for some time prior to suspicion. However, there are two alternative explanations for our findings. (1), it may be that phenotypes are ascertained in the EHR but not coded with ICDs. (2) it is possible that phenotypes developed over the course of the diagnostic process due to age related expressivity.

To examine these possibilities, we first looked at the timing of orders for transthoracic echocardiogram and ophthalmology exams -- two procedures used to ascertain key HCTD features – and complete blood counts (CBC) as a procedure that is unrelated to HCTD. Among the 87 HCTD patients, 67 (89%), 47 (54%), and 53 (61%) received a transthoracic echocardiogram, ophthalmology exam, and complete blood count, respectively. However, only a minority of transthoracic echocardiograms (9%) and ophthalmology exams (17%) occurred prior to suspicion, in contrast with complete blood counts (one of the most common clinical investigations), where 51% occurred prior to suspicion. (Table 4).

**Table 4:** A table demonstrating: A) the total the number of individuals to undergo relevant clinical investigations (Echocardiography, ophthalmologic exam) vs a control investigation (Complete blood count) prior to clinical suspicion and B) the total the number of investigations prior to clinical suspicion.

| Investigation | Individuals with investigation | Individuals (pre-suspicion) | Total numbers of investigations | Exams (pre-suspicion) |
| --- | --- | --- | --- | --- |
| Echocardiography, transthoracic | 67 (89%) | 16 (18%) | 308 | 28 (9%) |
| Ophthalmological exam | 47 (54%) | 15 (17%) | 278 | 46 (17%) |
| Complete blood count | 53 (61%) | 32 (37%) | 316 | 160 (51%) |

To rule out age-related expressivity and phenotyping error creating phenotypic convergence we manually reviewed patients with aortic aneurysms (AA) and ectasias that were ascertained via phecodes after clinical suspicion to ensure they were not present before suspicion but code for after and had not developed in between suspicion and diagnosis (n=24). All AA diagnoses were made with transthoracic echocardiograms that were present as a radiology report in the record. The first mention of AA occurred in the same time interval as the first ICD code in 20 of 24 cases (83%). These AAs were first discovered from an echocardiogram ordered in response to suspicion or diagnosis of an HCTD. One patient had a past echocardiogram with a noted mild dilation of the aorta that was not billed for nor discussed in the clinical notes; on repeat echo due to clinical suspicion of HCTD, the aneurysm had grown and a diagnosis code appeared in the record. In three cases (13%), an aortic aneurysm was noted just prior to suspicion, but the ICD code was not assigned in the post-suspicion era.

## Discussion

The clinical presentation of genetic disease has been studied extensively, but less attention has been paid to the way these diseases are reflected within EHR data. Understanding the phenotypic expression in the EHR is key to developing algorithms to detect undiagnosed patients. In this study, we develop a conceptual model to quantify data availability and the phenotypic manifestations of genetic disease from the perspective of the EHR. We demonstrate the relevancy of EHR fragmentation and ascertainment bias by applying our model to real world EHR data from a single academic health system for patients with genetically confirmed HCTDs. Analyzing the phenotypic signal throughout the ascertainment process suggested that many of the key phenotype’s indicative of an HCTD were ascertained only after a clinician suspected the disease. Thus, the EHR phenotypes of patients come to resemble the classical picture of the disease during the workup for and diagnosis of the HCTD in a phenomenon we named phenotypic convergence.

We found significant EHR fragmentation among our HCTD cohort. Only 34% of patients had EHR data for their full diagnostic trajectories – including visits prior to first suspicion through diagnostic confirmation. Many HCTD patients first came to our institution with a diagnosis or were referred to the hospital due to clinical suspicion. Of the 87 patients with fully ascertained trajectories, we found a wide range of ages of age at diagnosis, with 34% diagnosed as adults and 15% diagnosed after age 40, which suggests a wide window of opportunity to diagnose these patients at an earlier age.

We found that PheRS effectively distinguished between those the HCTD versus unaffected controls. However, the majority of HCTD features were ascertained after clinical suspicion and diagnosis. When using all available data, 34% of patients had highly elevated PheRS; when only data prior to suspicion was used, this number fell to 7%. The PheRS increased after every event in the diagnostic trajectory resulting in a gradual convergence on the classical phenotype known in the literature, a phenomenon we call phenotypic convergence. Even phenotypic attributes that were likely to have been present since birth (e.g congenital heart defects) or represent long term chronic consequences of HCTD (e.g. scoliosis) are often mentioned only after clinical suspicion for an HCTD diagnosis.

A deeper look at the diagnostic process revealed evidence that phenotypic convergence is driven by the diagnostic process itself. Clinical suspicion prompts the clinician to look for additional signs of the disease, and diagnosis leads to heightened surveillance for particular phenotypes. Both transthoracic echocardiograms and ophthalmology exams were far more likely to occur after clinical suspicion than before while CBC, a test unrelated to HCTD, were just as likely to be ordered before suspicion as after. Most AAs were discovered via a transthoracic echocardiogram that was ordered due to suspicion for HCTD. In other words, in most cases, the AA did not trigger clinical suspicion for HCTD; rather the clinical suspicion triggered the ascertainment of the AA.

Our findings have multiple implications to those who seek to develop algorithms that detect undiagnosed genetic disease. Firstly, we demonstrate that EHR fragmentation can be a barrier to studying pre-suspicion phenotypes in EHR data. Secondly, we demonstrate the importance of censoring data prior to suspicion to avoid data leakage wherein information present only after the prediction timepoint is used in the predictive model. Our results suggest that the concern of leakage is not merely theoretical. The diagnostic process itself elicits key phenotypes so that using data after suspicion may bias algorithm performance. Our conceptual model may be useful in preparing datasets to train or test algorithms that seek to identify undiagnosed patients.

Our work has some limitations. First, we only included genetically confirmed cases of HCTD, which reduced our sample size and potentially biased our dataset. Secondly our cohort is based on a single tertiary medical center based in a major metropolitan area with primary and secondary care inpatient and outpatient facilities. This prevents us being able to conclude if phenotypic convergence is a global phenomenon or one restricted to a particular medical context. Thirdly, the phenotypic analysis relied on ICD billing codes and may have missed phenotypes that were documented in the clinical notes. A limited manual chart review of aortic aneurysm revealed that ICDs were an accurate source of phenotypic information. Future work may assess a wider variety of phenotypes to determine if concepts in the narrative text, which can be extracted with natural language processing (NLP), increase the positive predictive value of algorithms. Algorithms that use ICDs have the advantage of portability, and sophisticated NLP pipelines may be difficult to implement in smaller community clinics. It would be valuable to assess which sources of phenotypic information (e.g. ICDs, clinic notes, problem lists, laboratory measurements) provided the greatest benefit, so that tradeoffs between portability and accuracy could be better assessed. Fourthly, we based our analysis on a single class of genetic diseases. Therefore, future work is needed to establish whether phenotypic convergence is a phenomenon seen in other genetic or non-genetic diseases.

## Conclusion

Here we present a conceptual model to evaluate the diagnostic trajectory of Mendelian genetic diseases in EHR data. Using this model, we found that the majority of EHRs for HCTD individuals were fragmented. We also observed that characteristic phenotypes for HCTD were more likely to be ascertained after clinical suspicion and diagnosis. We name this novel phenomenon phenotypic convergence. Our work can aid in the identification of diseases that could benefit from algorithmic diagnosis as we seek to shorten diagnostic delay.

## Data Availability

All data produced in the present study are available upon reasonable request to the authors

## Supplement

Table S1 – Gene List

Table S2 – PhecodeX features of HCTD

Table S3 – Median PheRS scores for HCTD and Wilcoxan-rank sum comparison with controls

Table S4 – Statistical comparison of PheRS between different diagnostic intervals

Table S5 – Percentage of high scoring individuals in different diagnostic intervals

Table S6 – Phenotypic features of MFS over time

Table S7 – Phenotypic features of EDS over time

Table S8 - Phenotypic features of LDS over time

Table S9 - Phenotypic features of STL over time

